# Structural disconnectivity from quantitative susceptibility mapping rim+ lesions is related to disability in people with multiple sclerosis

**DOI:** 10.1101/2020.12.10.20244939

**Authors:** Ceren Tozlu, Keith Jamison, Thanh Nguyen, Nicole Zinger, Ulrike Kaunzner, Sneha Pandya, Yi Wang, Susan A. Gauthier, Amy Kuceyeski

## Abstract

**Background:** Multiple Sclerosis (MS) is a disease characterized by inflammation, demyelination, and/or axonal loss that disrupts white matter pathways that constitute the brain’s structural connectivity network. Individual disease burden and disability in patients with MS (pwMS) varies widely across the population, possibly due to heterogeneity of lesion location, size and subsequent disruption of the structural connectome. Chronic active MS lesions, which have a hyperintense rim (rim+) on Quantitative Susceptibility Mapping (QSM) and a rim of iron-laden inflammatory cells, have been shown to be particularly detrimental to tissue concentration causing greater myelin damage compared to chronic silent MS lesions. How these rim+ lesions differentially impact structural connectivity and subsequently influence disability has not yet been explored.

**Objective:** We characterize differences in the spatial location and structural disconnectivity patterns of rim+ lesions compared to rim-lesions. We test the hypothesis that rim+ lesions’ disruption to the structural connectome are more predictive of disability compared to rim-lesions’ disruption to the structural connectome. Finally, we quantify the most important regional structural connectome disruptions for disability prediction in pwMS.

**Methods:** Ninety-six pwMS were included in our study (age: 40.25 ± 10.14, 67% female). Disability was assessed using Extended Disability Status Score (EDSS); thirty-seven pwMS had disability (EDSS ≥ 2). Regional structural disconnectivity patterns due to rim- and rim+ lesions were estimated using the Network Modification (NeMo) Tool. For each gray matter region, the NeMo Tool calculates a Change in Connectivity (ChaCo) score, i.e. the percent of connecting streamlines also passing through a lesion. Adaptive Boosting (ADA) classifiers were constructed based on demographics and the two sets of ChaCo scores (from rim+ and rim- lesions); performance was compared across the two models using the area under ROC curve (AUC). Finally, the importance of structural disconnectivity in each brain region in the disability prediction models was determined.

**Results:** Rim+ lesions were much larger and tended to be more periventricular than rim- lesions. The model based on rim+ lesion structural disconnectivity measures had better disability classification performance (AUC = 0.67) than the model based on rim- lesion structural disconnectivity (AUC = 0.63). Structural disconnectivity, from both rim+ and rim- lesions, in the left thalamus and left cerebellum were most important for classifying pwMS into disability categories.

**Conclusion:** Our findings suggest that, independent of the evidence that they have more damaging pathology, rim+ lesions also may be more influential on disability through their disruptions to the structural connectome. Furthermore, lesions of any type in the left cerebellum and left thalamus were especially important in classifying disability in pwMS. This analysis provides a deeper understanding of how lesion location/size and resulting disruption to the structural connectome can contribute to MS-related disability.

## INTRODUCTION

Multiple Sclerosis (MS) is a chronic disease characterized by inflammatory and demyelinating plaques within the central nervous system (CNS) (Weinshenker et al., 1991). Disability evolution is highly heterogeneous between people with MS (pwMS), making future prediction of disability progression very difficult (Barkhof, 2002; Pérez-Miralles et al., 2013). Conventional magnetic resonance imaging (MRI) techniques are highly sensitive in detecting white matter lesions in pwMS, however, the correlation between lesion load measured with T2 imaging and clinical impairment is still modest. This mismatch between traditional imaging biomarkers and clinical symptoms is known as the clinico-radiological paradox in MS (Barkhof, 2002; D. K. B. Li et al., 2006). Therefore, advanced imaging techniques like Quantitative Susceptibility Mapping (QSM) (De Rochefort et al., 2010; Deh et al., 2015) may provide more information about lesion pathology (Wisnieff et al., 2015), that, in turn, may improve the understanding of the clinical implications of MS lesions (Chen et al., 2014; Kaunzner et al., 2019; Yao et al., 2018; Zhang et al., 2019).

QSM has been found to be more sensitive than conventional T2, T2^*^, and R2^*^ in the detection of iron accumulation in both gray and white matter regions in the brain (Cronin et al., 2016; Deistung et al., 2013; Langkammer et al., 2013; Stüber et al., 2016). Iron concentration in deep gray matter structures (thalamus and globus pallidus) (Zivadinov et al., 2018) has been shown to be significantly correlated with disability in pwMS; iron accumulation has also been identified in some white matter lesions in pwMS (Zhang et al., 2019). Furthermore, white matter lesions with a hyperintense rim appearance on QSM (QSM rim+ lesions) have increased inflammation on PK11195-PET, a finding which was histopathologically confirmed by the presence of inflammatory cells in the rim, as well as larger volume and more myelin damage compared to QSM rim- lesions (Kaunzner et al., 2019; Yao et al., 2018). Greater myelin damage in rim+ lesions has been shown (Gillen, Mubarak, Nguyen, & Pitt, 2018), and, furthermore, paramagnetic rim lesions as identified on susceptibility-based MRI have been cross-sectionally associated with worse disability (Absinta et al., 2019). However, there is no larger study to date that has demonstrated the relationship of disability with rim lesions that are identified with QSM in MS.

In addition to the differential pathology of the lesion, the clinical impact of a lesion is also related to its size and location and subsequent impact on the wider structural and functional connectivity networks. In the past decades, structural and functional connectome disruptions have been related to motor and cognitive dysfunction and depression in pwMS (Ceccarelli et al., 2010; Dineen et al., 2009; He et al., 2009; A. Kuceyeski et al., 2018; Y. Li et al., 2013; Llufriu et al., 2012; Nigro et al., 2015; Pagani et al., 2019). One way to investigate structural network disruptions of lesions is with the Network Modification (NeMo) Tool (A. Kuceyeski, Maruta, Relkin, & Raj, 2013). The NeMo Tool uses a database of healthy tractograms on which the MS-related lesion masks are super-imposed to estimate the resulting regional disconnectivity pattern. This approach has been used by our group and others to relate lesion-related structural disconnectivity patterns to impairments, outcomes, functional connectivity disruptions, rehabilitation response and gray matter pathology in pwMS (T. A. Fuchs et al., 2018; Tom A. Fuchs et al., 2018, 2020; A. F. Kuceyeski et al., 2015; A. Kuceyeski et al., 2018).

In this paper, we aim to characterize the differential impact of QSM rim- and rim+ lesions on the structural connectome, and, furthermore to test the hypothesis that rim+ lesions’ disruption to the structural connectome are more predictive of disability. To do this, we compared prediction accuracies of models classifying pwMS into disability categories using estimates of regional structural (white matter) connectome disruption due to rim- and rim+ lesions. If rim+ lesions are more impactful in terms of clinical disability through their disruption of the structural connectome, then models based on these measures should perform better than the rim- lesions’ structural connectome disruption patterns. A secondary aim of this work was to identify which brain regions’ structural disconnections are most important in the classification of pwMS into disability categories. It must be noted that this type of approach doesn’t consider the pathology type or severity of tissue damage within the lesion that may vary with lesion type – it only considers the lesion size and location and subsequent disruption of the structural connectivity network. If we can better understand how different lesion types can impact clinical outcomes through their disruption of the structural network, we may be able to better identify those patients at risk of disability and adjust treatments to minimize the burden of MS.

## MATERIAL AND METHODS

### Subjects

This is a retrospective study of a cohort of 96 pwMS (age: 40.2 ± 10.1, 67% females) with a diagnosis of CIS/MS (CIS = 8, RRMS = 87, PPMS = 1). Demographic data was collected (age, sex, race, treatment duration and disease duration), subjects underwent an MRI scan and Extended Disability Status Score (EDSS) was used to quantify disability. PwMS were categorized into two groups: those with no disability (EDSS < 2) or those with disability (EDSS ≥ 2). This classification is based on EDSS values of 0-1.5 representing some abnormal signs but no functional disability appreciated. All studies were approved by an ethical standards committee on human experimentation, and written informed consent was obtained from all patients.

### MRI data acquisition and Processing

MRIs were acquired using a 3T GE scanner (Hdxt 16.0) with an 8-channel phased-array coil. Anatomical T1-weighted sagittal 3D-BRAVO (1.2×1.2×1.2mm), T2 (0.5×0.5×3mm), T2-FLAIR (1.2×0.6×0.6mm) sequences were acquired. A QSM image was reconstructed from complex GRE images (TR = 57 ms, first TE = 4.3 ms, echo spacing = 4.8 ms, echo train length = 11, axial FOV = 24 cm, phase FOV factor = 0.8, acquisition matrix = 416×320 interpolated to 512×512, slice thickness = 3 mm, flip angle = 20°, bandwidth = 244 kHz, number of signal averages = 0.75, readout bandwidth = ±62.5 kHz) using a fully automated Morphology Enabled Dipole Inversion (MEDI+0) method zero-referenced to the ventricular cerebrospinal fluid (Liu, Spincemaille, Yao, Zhang, & Wang, 2018; Spincemaille et al., 2019).

The conventional images (T1w, T2w, T2w FLAIR) was co-registered to the GRE magnitude images using the FMRIB’s Linear Image Registration Tool algorithm (Jenkinson, Bannister, Brady, & Smith, 2002); automated brain segmentation was performed using FreeSurfer (Fischl, Sereno, & Dale, 1999). White matter (WM) and gray matter (GM) masks were manually edited for misclassification due to WM T1-hypointensities associated with lesions. The WM hyperintensity lesion masks were created from the T2 FLAIR images by categorizing the tissue type based on the image intensities within the Lesion Segmentation Tool (LST) and were further edited if necessary. Next, the T2FLAIR lesions masks were coregistered to the QSM images and further edited (if needed) to better match the lesion geometry on QSM. The presence of a hyperintense QSM rim was determined by trained neurologist (U.K.) and neuroradiologist (W.H.). In the case of disagreement of two reviewers, an independent third neurologist (S.G.) decided on the presence of a positive hyperintense rim. Once the rim+ lesions were identified, they were removed from the T2FLAIR lesion masks to obtain a rim- lesion mask.

QSM rim+ lesion masks were transformed to the individual’s T1 native space using the inverse of the T1 to GRE transform and nearest neighbor interpolation. Individual T1 images were then normalized to MNI space using FSL’s linear (FLIRT) and non-linear (FNIRT) transformation tools (http://www.fmrib.ox.ac.uk/fsl/index.html); transformations with nearest neighbor interpolation were then applied to transform both native anatomical space lesion masks to MNI space. Lesions were manually inspected after the transformation to MNI space to verify accuracy. The MNI space rim- and rim+ lesion masks were processed through the newest version of the Network Modification (NeMo) Tool (Amy Kuceyeski, Maruta, Relkin, & Raj, 2013), NeMo Tool 2.0, that estimates the resulting pattern of structural disconnectivity due to a given lesion mask. NeMo Tool 2.0 calculates the Change in Connectivity (ChaCo) score for each of 86 cortical, subcortical and cerebellar regions, which is defined as the percent of tractography streamlines connecting to that region that also pass through the lesion mask. The newest version of the tractography database consists of structural connectomes from 420 unrelated healthy controls (206 female, 214 male, 28.7 ± 3.7 years), see Supplemental Information for details on the creation of the tractography database. ChaCo scores were extracted separately from rim- and rim+ lesion masks. The ChaCo scores from the rim- lesion mask were computed across all subjects, while the ChaCo scores from rim+ lesion masks were computed only for the subjects who had at least one rim+ lesion (N=56). The ChaCo scores for the subjects without rim+ lesions were 0, since there was no structural disconnectivity due to rim+ lesions for these subjects. To test for differences in the regional disconnectivity patterns of rim+ and rim- lesions, a Wilcoxon rank-sum test was performed on the regional ChaCo scores from the rim- and the rim+ lesion masks over the 56 pwMS that had at least one rim+ lesion. A Wilcoxon rank-sum test was used to test for differences in regional structural disconnectivity between disability groups for both rim+ and rim- lesion ChaCo scores. Regions were considered significantly different if p<0.05, after Benjamini-Hochberg (Benjamini & Hochberg, 1995) correction for multiple comparisons.

### Modeling and statistical analysis

Classification was performed using the ADA method, a boosting algorithm of decision trees (Alfaro, Gáamez, & García, 2013), see Supplementary Material for details. For the classification task into disability and no disability groups, three models were created based on demographic/clinical variables (age, sex, race, disease duration and treatment duration) and i) ChaCo scores from rim- lesions (Model I), ii) ChaCo scores from the rim+ lesions (Model II), iii) both sets of ChaCo scores from rim- and rim+ lesions (Model III).

The ADA model was trained with two cross-validation loops to optimize the hyperparameters and build the model (5-fold inner) and test the performance on hold-out data (5-fold outer), see Figure 1. The inner loop performed grid-search to find the set of hyperparameters that maximized area under the Receiver Operating Characteristics curve (AUC) in the validation set. Synthetic Majority Over-sampling Technique (SMOTE) (Chawla, Bowyer, Hall, & Kegelmeyer, 2002) was used to obtain a class-balanced training dataset to improve the prediction accuracy for the minority class. SMOTE compensates for imbalanced classes by creating synthetic examples using nearest neighbor information instead of creating copies from the minority class, and has been shown to be among the most robust and accurate methods with which to control for imbalanced data (Santos, Soares, Abreu, Araujo, & Santos, 2018). The inputs are standardized in the inner loop and in the outer loop to avoid data-leakage. A final model built using the entire training dataset with the optimal hyperparameters and assessed on the hold-out test set from the outer loop. The outer loop was repeated using 100 different random partitions of the data. The average of AUC (over all 5 folds x 100 iterations = 500 test sets) were calculated to assess the performance of the models. Performance metrics for the three models were compared with the Kruskal-Wallis and Wilcoxon rank sum test. The models were considered significantly different when p < 0.05 (after Benjamini-Hochberg correction for multiple comparisons). The relative importance of the input variables in the final ADA models was calculated using the weight of the tree and gain of the Gini Index, which is given by a variable in a tree (Alfaro et al., 2013). The software R (https://www.r-project.org) version 3.4.4 and Matlab version R.2020a were used for all statistical analyses and graphs.

**Figure 1:**
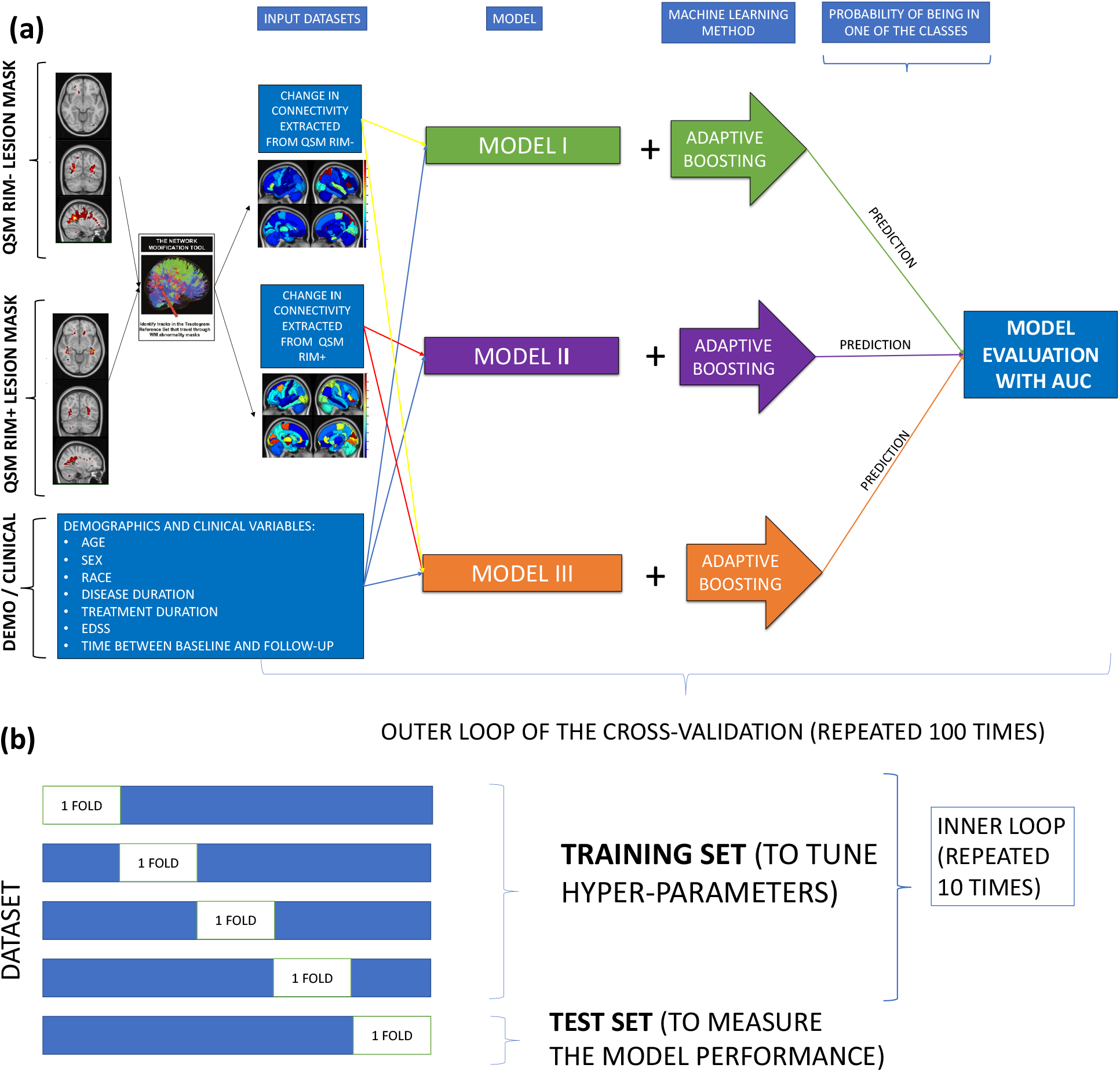
Overview of the analytic pipeline workflow, including (a) calculation of ChaCo scores from rim- (T2 FLAIR lesions without rim+ lesions) and rim+ lesion masks and ADA models, and (b) the cross-validation scheme for the training and testing of the ADA models.

## RESULTS

### Characteristics of pwMS

Table 1 displays subject demographics, clinical information and the number of rim- and rim+ lesions. PwMS that had no disability (N = 59) were significantly younger, had lower disease duration and significantly fewer rim- lesions than those with disability (N = 37). A higher percent of those pwMS with at least one rim+ lesion had disability (45%) compared to the percent of pwMS with no rim+ lesions and disability (30%), see Supplementary Table 1. While rim+ lesions were less frequent across the population and less numerous within an individual, they were much larger in volume (mean 484 cc, IQR: [173, 572]) than rim- lesions (mean 141 cc, IQR: [69, 145]). Heat maps of lesion masks for the two lesion types are shown in Figure 2, where it can be appreciated that rim+ lesions tended to cluster in periventricular white matter, compared to rim- lesions that are more widespread throughout the white matter.

**Table 1.** Clinical, demographic and imaging characteristics for all 96 pwMS (first column) and split into disability categories (second column). Values are presented as median [1^st^ quartile, 3^rd^ quartile] for the continuous variables, p-values are corrected for multiple comparisons. Age, disease and treatment duration were measured in years. *indicates corrected p < 0.05.

|  | **All (n=96)** | **No disability (n=59) vs disability (n=37)** |
| --- | --- | --- |
| **Female%** | 67 | 68 vs 65 (p=0.94) |
| **Race %** | African American: 16<br>Asian: 2<br>Caucasian: 72<br>Hispanic: 5<br>Other: 5 | African American: 11 vs 24<br>Asian: 1 vs 2<br>Caucasian: 76 vs 64<br>Hispanic: 5 vs 2<br>Other: 5 vs 5<br>(p=0.56) |
| **Age** | 38 [32, 48] | 35 [30, 42] vs 47 [37, 51]* (p<0.05) |
| **Disease duration** | 4.6 [2.5, 11.2] | 4.1 [2.2, 8.3] vs 7.9 [3.2, 15.9]* (p<0.05) |
| **Treatment duration** | 3.1 [1.5, 6.1] | 2.4 [1.3, 4.4] vs 3.7 [2.3, 8.0] (p=0.06) |
| **EDSS** | 1 [0, 2] | 0 [0, 1] vs 2.5 [2, 4]* (p<0.05) |
| **# of rim- lesions** | 22 [11, 57] | 18 [8, 35] vs 42 [15, 62]* (p<0.05) |
| **# of rim+ lesions** | 1 [0, 4] | 1 [0, 3] vs 2 [0, 5] (p=0.15) |

**Figure 2:**
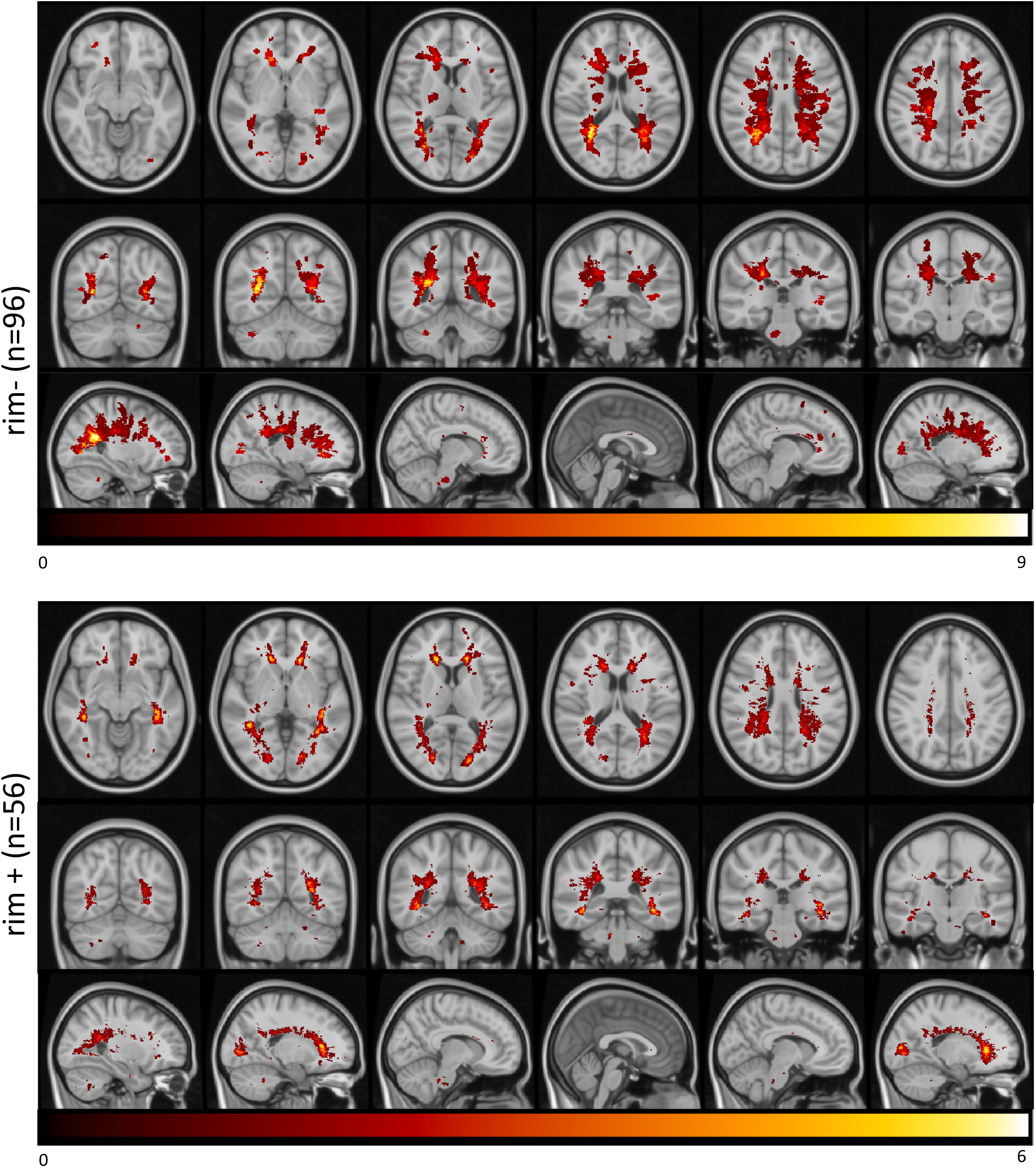
Heatmap of the lesion masks extracted from rim- (T2 FLAIR lesions without rim+ lesions, first row) and rim+ (second row) images. Color indicates the number of individuals (out of 91) that had a lesion in that voxel.

### Structural disconnectivity from rim- and rim+ lesions

Regional ChaCo scores of structural disconnectivity based on the rim- and rim+ lesion masks are visualized in Figure 3. Median ChaCo scores from rim+ lesion masks were computed only for the subjects who had rim+ lesions (N=56), while the median ChaCo scores from the rim- lesion mask was computed across all subjects. Note the scale differences in the two modalities – this is mostly due to the fact that there were far fewer rim+ lesions than rim- lesions. Left paracentral, left precuneus, and bilateral precentral (primary motor) regions had high disconnectivity in both the rim- and rim+ lesion masks. Right and left putamen also had relatively high disconnectivity from the rim- and rim+ lesion masks, respectively. ChaCo scores from rim- lesion masks were significantly higher than those from rim+ lesion mask for all regions (p<0.05, corrected), particularly in right primary motor and right paracentral gyrus.

**Figure 3:**
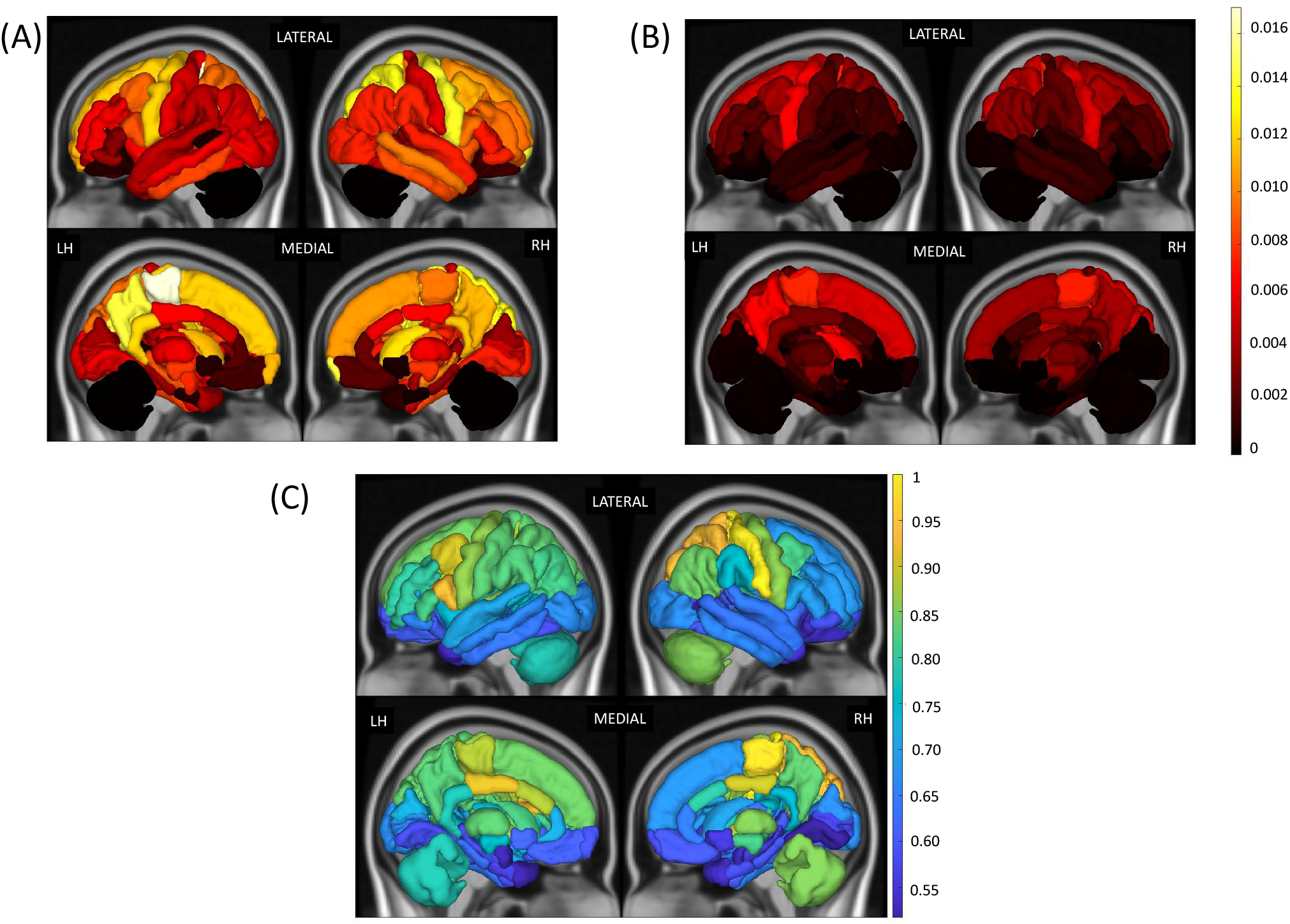
Median of ChaCo scores extracted from (A) rim- lesion mask (T2 FLAIR lesions excluding rim+ lesions) across all pwMS (N=96) and (B) rim+ lesion masks, only for the pwMS who had at least one rim+ lesion (N=56). (C) Relative paired Wilcoxon rank-sum statistic indicating all regions had greater structural disconnection from rim- lesion masks than from rim+ lesion masks (considering only the 56 pwMS who had at least one rim+ lesion).

### Structural disconnectivity differences across disability subgroups

Figure 4 illustrates the median Chaco scores for each subgroup of categories, i.e., no disability vs with disability for the two lesion types. The ChaCo scores based on rim- lesion masks were significantly larger in 22 regions, most prominently in the left frontal areas, in the pwMS with disability compared to those without disability (p<0.05, corrected), see Supplementary Figure 1. There were no significant differences between the disability subgroups for the ChaCo scores based on rim+ lesion masks.

**Figure 4:**
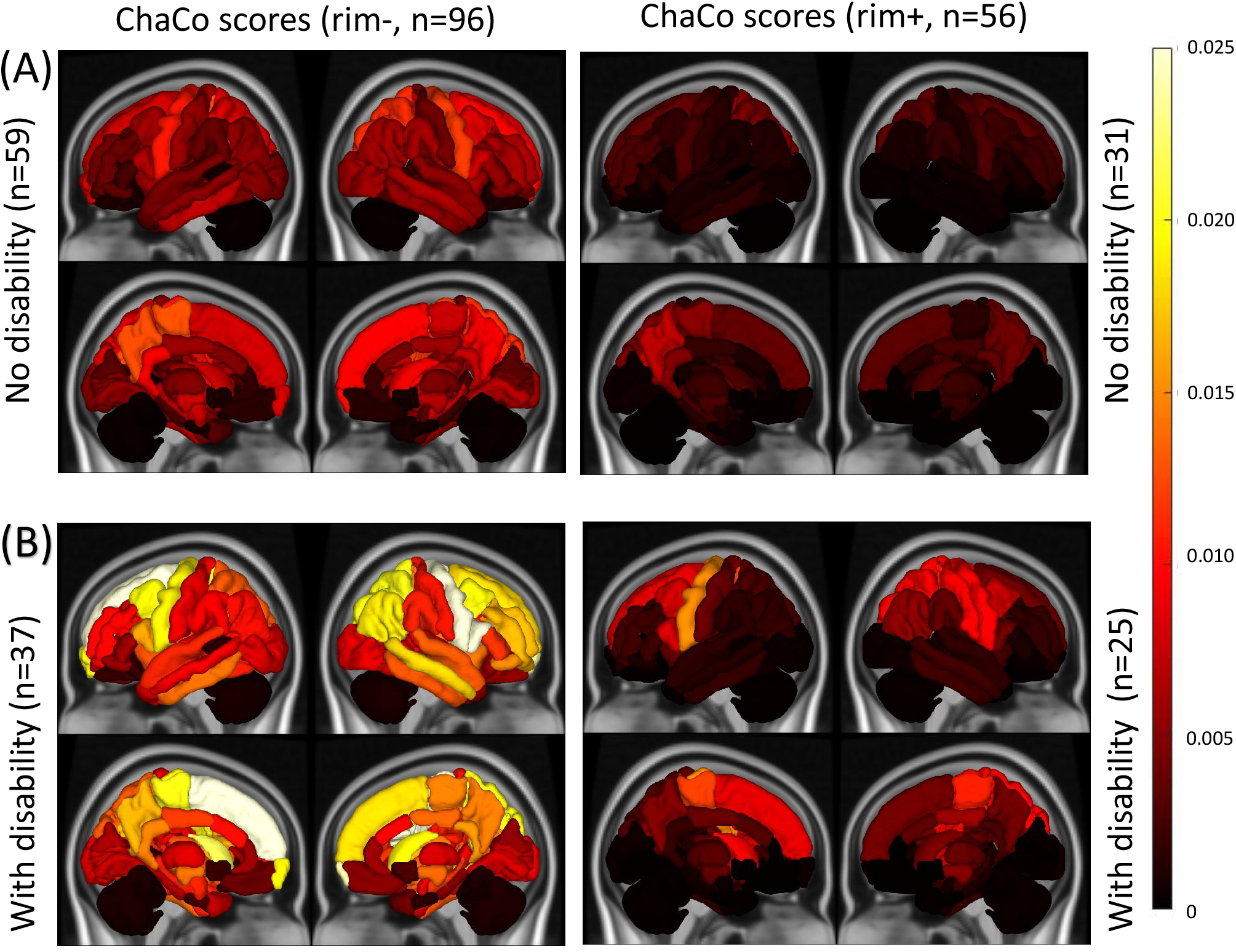
Median of ChaCo scores extracted from rim- (T2 FLAIR lesions excluding rim+ lesions) and rim+ lesion masks for pwMS (A) no disability vs (B) those with disability.

### Classification results

Figure 5 depicts the three models’ distributions of AUC over the 100 outer loops and 5 test datasets for each outer loop for the disability classification task. Model II, which included demographics/clinical variables and ChaCo scores from the rim+ lesions, had significantly higher AUC than the other two models. Supplementary Figure 2 illustrates balanced accuracy, sensitivity, and specificity for the three models for comparison to previous findings.

**Figure 5:**
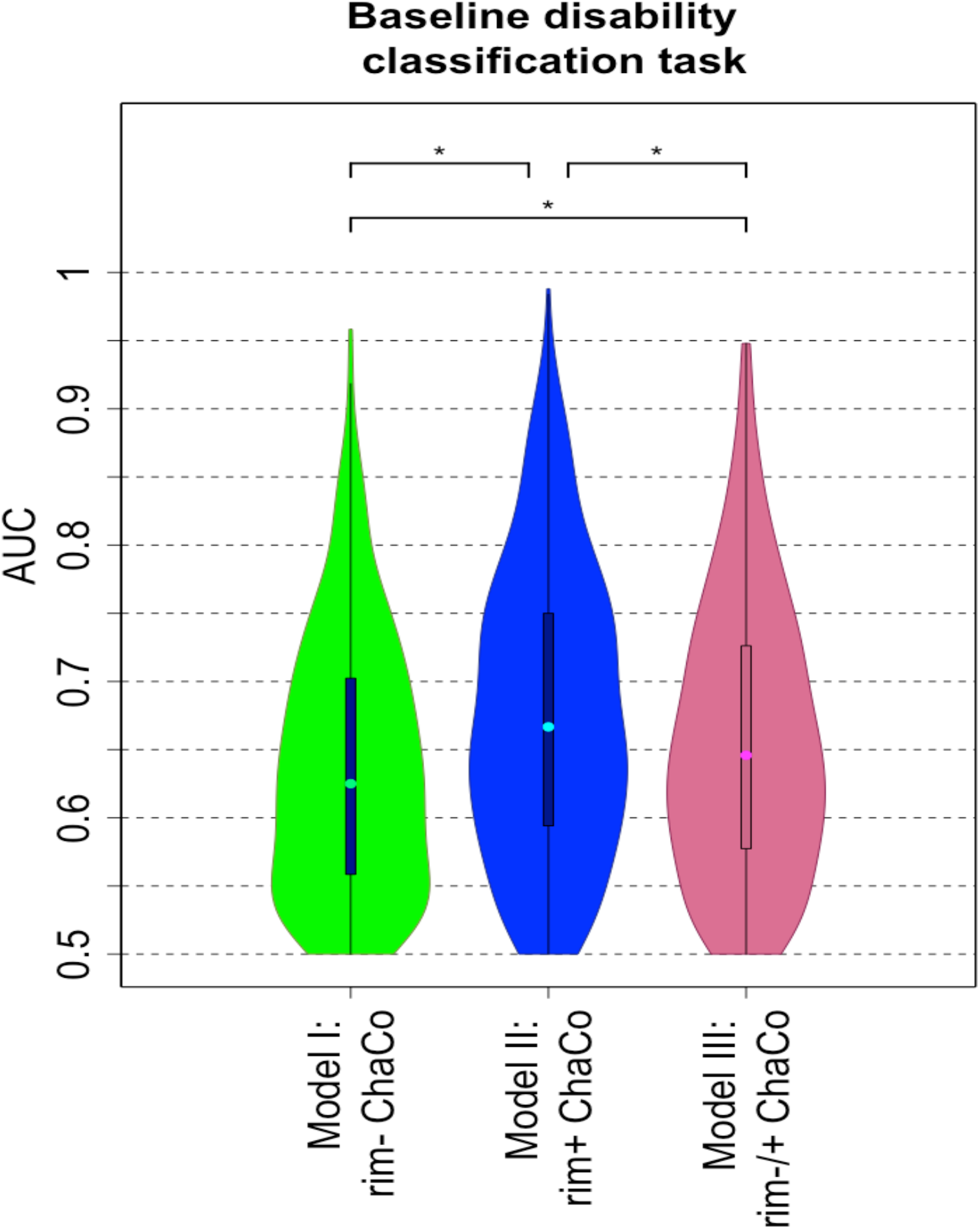
AUC results obtained with Model I (structural disconnectivity from rim- lesion masks), Model II (structural disconnectivity from rim+ lesion masks), and Model III (both rim- and rim+ lesion structural disconnectivity) for the classification task of disability vs no disability. *indicates significant differences in AUC, corrected p < 0.05.

### Variable Importance

Figure 6 shows the feature importance of the structural disconnectivity measures (ChaCo scores) and demographics/clinical variables for Models I and II in classifying disability. The third quartiles of the feature importance scores were visualized since the data was highly skewed. Structural disconnection in the left cerebellum and left thalamus due to both rim- and rim+ lesions were the most important variables in classifying disability. Race and disease duration were the most important demographics/clinical variables in Model I and II, respectively.

**Figure 6:**
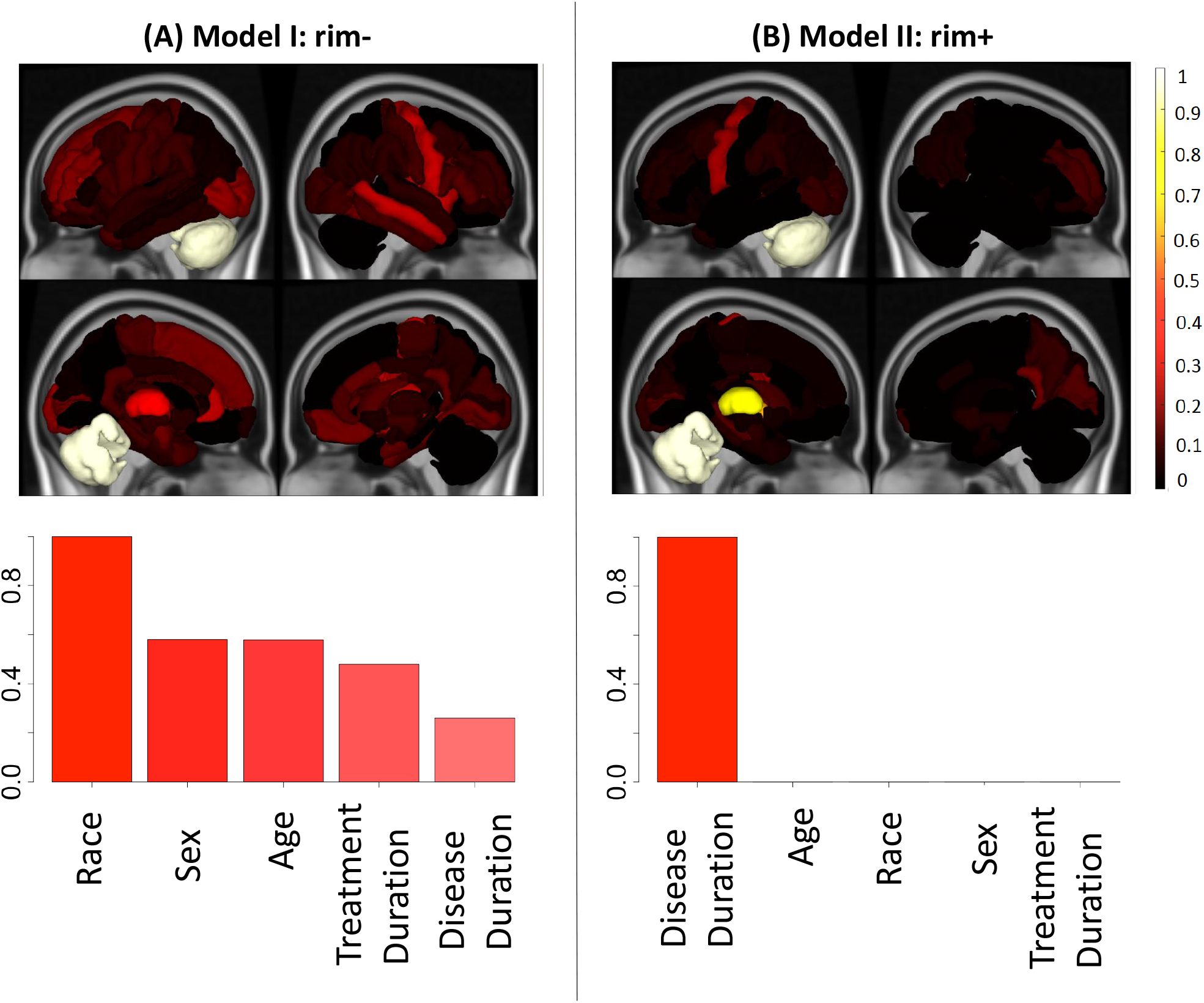
Relative feature importance for the models that included demographics and regional structural disconnectivity (ChaCo scores) due to (A) Model I: rim- lesions (T2 FLAIR lesions excluding rim+ lesions) (left column) and (B) Model II: rim+ lesions (right column) for the classification of pwMS with disability vs those with no disability. Feature importance for the regional ChaCo scores are visualized via brain volumes and demographic variable importance by bar plots. Third quantiles of the feature importance distributions are visualized due to the distribution skewness. Relative importance values for all figures were obtained by dividing that variable’s feature importance by the maximum importance value across both models.

## DISCUSSION

In this study, we investigated the patterns of structural connectome disruption arising from two types of lesions that occur in pwMS, namely those with a hyperintense rim on QSM (rim+) and those without. We compared the classification accuracy of models using the structural disconnectivity of the two lesion types to predict disability categories in pwMS. Our main findings were that 1) pwMS had high structural disconnectivity in motor regions (precentral and paracentral gyri) resulting from both types of lesions, 2) rim+ lesions were larger than rim- lesions, tended to more frequently occur in periventricular areas and thus impact structural connectivity disproportionately in periventricular regions, 3) structural disconnectivity from rim+ lesions better classified pwMS into disability categories than structural disconnectivity from rim- lesions, and 4) structural disconnectivity in left cerebellum and left thalamus resulting from both lesion types were among the most important features in the disability classification.

### Structural disconnectivity from MS lesions is highest in motor regions and rim+ lesions are more periventricular than rim- lesions

The regions with the highest structural disconnection scores from both types of lesions were mostly motor-related, including paracentral, precentral and putamen. It appears from Figure 2 and 3 that, in addition to the rim- lesions being more numerous and thus having larger disconnectivity measures, the two lesion types tend to have slightly different spatial locations and regional patterns of structural disconnectivity. The rim- lesions appear distributed widely throughout the white matter while the rim+ lesions tend to cluster around the ventricles. The ChaCo scores reflect this in that most of the regions with disconnection from the rim+ lesions are indeed periventricular and the remainder of the brain is relatively spared; this stands in contrast to the ChaCo scores from rim- lesions that are more widespread across the brain.

### Structural disconnectivity from rim+ lesions better predicts disability in pwMS

Previous cross-sectional studies classifying pwMS into disability categories based on imaging biomarkers, including connectome measures, have shown similar prediction performance with an accuracy and AUC between 0.50 and 0.67 (Zhong et al., 2017; Zurita et al., 2018). The relationship between connectome disruption due to T2FLAIR lesions and cross-sectional and longitudinal disability change, such as processing speed deficits, has been previously assessed (A. F. Kuceyeski et al., 2015; A. Kuceyeski et al., 2018). Our findings suggest that, independent of evidence they have more damaging pathology, rim+ lesions also may be more influential on disability through their disruptions to the structural connectome. We conjecture that structural disconnectivity due to rim+ lesions may be more detrimental due to their 1) much larger volume, as seen in this and other work (Zhang et al., 2019), and 2) periventricular location, which may result in damage to white matter that is more central in the brain and thus more disruptive to the overall network structure. It must be emphasized that only the structural disconnectivity due to the lesion, which is influenced by its size and location, was used in our classification models of disability. No information about the pathology, type or severity of tissue damage within the lesion (such as iron concentration, demyelination, axonal loss, presence of inflammation, edema, etc.) was captured in our study. Therefore, if rim+ lesions have more severe pathologies and thus more severe damage to structural connections, for example, we would not be capturing that in our ChaCo scores. Thus, our current results show that, independent of lesion pathology, rim+ lesion size/location and subsequent structural disconnection may have a greater impact on disability than rim- lesions in pwMS.

PwMS with disability had significantly more rim- lesions than those without disability, while there were no differences in the number of rim+ lesions between the disability groups. However, the model based on structural disconnection from rim+ lesions better predicted disability than the one based on structural disconnection from rim- lesions. This result supports the notion that disability does not directly track with the number of rim+ lesions, but rather, more likely, the size and/or location of the lesions and how they can disrupt structural connectivity networks.

### Disconnection in the left thalamus and left cerebellum are central to accurate disability classification

Examination of the feature importance values from the classification models revealed the central role of structural disconnectivity in the left thalamus and left cerebellum. Interestingly, these same regions were important regardless of the lesion type (rim+ or rim-) causing disconnectivity. The thalamus plays an important role in a wide range of functions such as cognition, memory, executive function, and motor ability (Alexander, DeLong, & Strick, 1986; Batista et al., 2012; Henry et al., 2008) and is known to be among the most affected regions in pwMS (Vercellino et al., 2009). Functional connectivity changes and structural changes in the thalamus, observed with anatomical and diffusion MRI, have been related to cognitive and motor impairment (Henry et al., 2008; Schoonheim et al., 2015; Tovar-Moll et al., 2009). Previous work has shown the thalamus to be one of the only regions exhibiting a significant relationship between atrophy and structural disconnection in pwMS, which indicates this region may be particularly vulnerable to increased atrophy when lesions occur in its connecting white matter (A. F. Kuceyeski et al., 2015). In addition, it has been shown that more thalamic atrophy was significantly related to increased EDSS in pwMS (Tao et al., 2009; Tovar-Moll et al., 2009) and that pwMS with disability had significantly lower thalamic volume compared to healthy controls, while no differences were found between non-impaired pwMS and controls (Zhong et al., 2017). Taken together, these studies indicate the central role of the thalamus in the development of disability in pwMS.

Many studies have shown relationships between cerebellar pathology and impairments in motor control and cognition (D’Ambrosio et al., 2017; Weier et al., 2014). The presence of cerebellum-related symptoms at the onset of MS such as coordination issues or tremor were i) shown to be associated with shorter time to an EDSS of 6 (Weinshenker et al., 1991) and ii) related to earlier onset of progressive disease diagnosis (Novotna et al., 2015). Atrophy in anterior cerebellum was associated with motor dysfunction (D’Ambrosio et al., 2017) and reduced total cerebellar volume was related to worse cognitive test performance (Weier et al., 2014) in pwMS.

### Race and sex play a potentially important role in disability classification

Race was one of the most important demographic predictors in the rim-model. It has been shown that African Americans (which make up the largest non-Caucasian group in our study) generally have larger T1 and T2 lesion volumes, more severe disability at diagnosis and more severe disease progression (Cipriani & Klein, 2019; B. Weinstock-Guttman et al., 2010; Bianca Weinstock-Guttman et al., 2003). Sex also appeared to be an important predictor in the rim- lesion model for disability classification; specifically, being male was associated with higher probability of being in the disability group. It has been shown previously that male patients tend to have more severe disease onset with accelerated clinical progression in MS (Bove et al., 2012; Debouverie, Pittion-Vouyovitch, Louis, & Guillemin, 2008; Gholipour, Healy, Baruch, Weiner, & Chitnis, 2011). Disease duration was also an important feature in the rim+ ChaCo model; this is unsurprising as EDSS generally increases over the course of the disease.

#### Limitations

The main limitation of our study is sample size; classifiers are always more robust and when they are trained and tested on larger datasets, particularly when dealing with such a heterogenous disease as MS. Another drawback was the use of the NeMo tool, which estimates a lesion’s structural disconnectivity based on a database of healthy controls that may not perfectly reflect the particular individual’s structural connectivity network. However, MS lesions do disrupt diffusion MRI signals and can add noise to tractography results, so the NeMo Tool may be a good alternative with which to estimate structural disconnection. Another limitation is that the models used in our study considered only the lesion size and location and their subsequent regional structural disconnection. A future study may consider the impact of the severity and type of tissue damage within the different lesions on structural disconnectivity and subsequent disability. Finally, this work only explored cross-sectional relationships; future work should work to predict probability of disease progression over time for use in clinical care decisions.

#### Conclusions

This work represents, to the best of our knowledge, the first to quantify and examine the differential impact of rim+ and rim- lesions on the structural connectome and, furthermore, to use these measures of disconnectivity to classify pwMS into disability categories. Structural disconnectivity associated with rim+ lesions on QSM was more related to disability than structural disconnectivity associated with rim- lesions. Damage the structural connections of the left cerebellum and thalamus from either lesion type were especially impactful on disability. This analysis provides a deeper understanding of how different lesion types can disrupt the structural connectome and contribute to MS-related disability. Deeper understanding of the role of the connectome in MS is needed if we are to gain a comprehensive view of the disease to ultimately improve clinical outcomes in pwMS.

## Supporting information

IRB approval document

## Data Availability

The structural disconnectivity scores that support the findings of this study are available upon reasonable request. The codes that were used in this study are available at https://github.com/cerent/MS-QSM

## Acknowledgements

This work was supported in part by the following grants: R21 NS104634 (AK), R01 NS102646 (AK), R01 NS104283 (SG), R01 NS090464 (YW), R01 NS105144 (SG, YW), and UL1 TR000456 (SG) from the Weill Cornell Clinical and Translational Science Center (CTSC). We would like to thank Eric Morris and Weiyuan Huang for their role in helping to edit, organize, and curate the clinical and imaging data.

## Author contribution

C.T. designed the study, helped with processing the MRI data, carried out the statistical and machine learning analyses, drafted and wrote the manuscript.

K.J. developed the version 2.0 of Network Modification tool and edited the manuscript.

N.Z. collected and merged data.

T.N. assisted with image data processing and edited the manuscript.

U.K. collected the data and edited the manuscript.

S.P. performed data processing and edited the manuscript.

Y.W. assisted with data acquisition and analyses and edited the manuscript.

S.G. collected the data, supervised the analyses, helped interpret results, and edited the manuscript.

A.K. designed the study, supervised the analyses, and edited the manuscript.

## Citation gender diversity statement

We used classification of gender based on the first names of the first and last authors (Dworkin et al., 2020), with possible combinations including male/male, male/female, female/male, and female/female. The gender balance of papers cited within this work was quantified using gender-api.com. The authors with a gender estimation accuracy lower than 90% were checked using manual gender determination from authors’ publicly available pronouns. Among the 55 cited works, 1 article had only one author. Among the 54 cited works with more than one author, %48 (n = 26) were MM, %24 (n = 13) were WM, %19 (n = 10) were MW, and %9 (n = 5) were WW.

## Abbreviations

ADA: adaptive boosting
AUC: area under curve
ChaCo: change in connectivity
EDSS: extended disability status score
GM: gray matter
QSM: quantitative susceptibility mapping
LST: lesion segmentation tool
MS: multiple sclerosis
NeMo: network modification
pwMS: people with multiple sclerosis
WM: white matter

