## Supplementary Materials for "Structural disconnectivity from quantitative susceptibility mapping rim+ lesions is related to disability in people with multiple sclerosis"

**Running title:** Disability classification in MS

Ceren Tozlu[1], Keith Jamison[1], Thanh Nguyen[1], Nicole Zinger[2], Ulrike Kaunzner[2], Sneha Pandya[1], Yi Wang[1], Susan A. Gauthier[2], Amy Kuceyeski[1,3]

[1]Department of Radiology, [2]Department of Neurology and [3]Brain and Mind Research Institute, Weill Cornell Medicine, New York, NY, USA

### Supplementary Material

#### NeMo Tool

The Network Modification 2.0 (NeMo 2.0) tool estimates structural connectivity disruption due to a lesion or injury from a database of healthy structural connectivity. We first computed a database of whole-brain tractograms for 420 unrelated subjects from the Human Connectome Project Young Adult (HCP-YA) dataset. The HCP diffusion data consists of 1.25mm isotropic voxels, 3 shells (b=1000,2000,3000) and 90 directions per shell, collected with both R-L and L-R phase encoding. HCP data have been minimally preprocessed to correct for motion, EPI and eddy-current distortion, and registered to subject T1 anatomy (M. F. Glasser et al., 2013). We used MRtrix3 to estimate a voxel-wise multi-shell, multi-tissue constrained spherical deconvolution (CSD) model (Jeurissen, Tournier, Dhollander, Connelly, & Sijbers, 2014), followed by whole-brain probabilistic tractography (iFOD2 (J Donald Tournier, Calamante, & Connelly, 2010) with anatomically constrained tractography – ACT (Smith, Tournier, Calamante, & Connelly, 2012)) using dynamic seeding to produce 5 million streamlines per subject. We also computed streamline weights to reduce known biases in tractography algorithms and better match the whole brain weighted tractogram to diffusion properties of the observed data (SIFT2, (Smith, Tournier, Calamante, & Connelly, 2015)). Streamlines for each HCP subject were warped into a common volumetric space (MNI152).

Given a lesion mask in MNI space, the NeMo tool identifies the gray matter endpoints of all streamlines that pass through the masked voxels, and computes a Change in Connectivity (ChaCo) score for each gray matter region in a given atlas, representing the fraction of streamlines connecting to that gray matter region that have passed through the lesion. Disconnectivity is connected for each region for each of the 420 HCP database subjects, and then averaged across database subjects to create the final group ChaCo estimate for each region. A region with a ChaCo score of 0 is expected to have normal structural connectivity, while a ChaCo score of 1 suggests complete disconnection.

**Adaptive Boosting method and the parameters used in this study**

Adaptive Boosting (ADA) consecutively applies decision trees that split the data into two classes successively based on a randomly chosen variable at each node. Decision trees in ADA chose the variable which minimizes the Gini Index (GI) in the classification analysis. GI at node *t* is defined as

$$\sum_{c=1}^{L} \widehat{p_{tc}}\,(1 - \widehat{p_{tc}})$$

where $\widehat{p_{tc}}$ is the proportion of the observation in class *c* at node *t*.

*adabag* library and *boosting* function in R version 3.4.4 were used for the ADA model creation. The minimum number of observations that must exist in a node in order for a split to be attempted (*minsplit*) and complexity parameter (*cp*) were optimized in the inner loop of cross-validation. The default value of *minsplit* and *cp* parameters are 20 and 0.01 in rpart.control function of R (Breiman, Friedman, J.Stone, & Olshen, 1984). To find the optimal value of these parameters for different train datasets, a grid search was applied for *minsplit* in the interval [5, 50] with steps of 5 and for *cp* in the interval [0.001, 0.1] by increments of 10. The pair of *minsplit* and *cp* parameters that maximized AUC in the inner loop was identified as the optimal pair of hyperparameters and then used to build the model. For each model, 50 decision trees were used.

**Supplementary Table 1:** Number of pwMS who had disability and no disability without rim+ lesion vs at least one rim+ lesion.

| | **No rim+ lesions** | **At least one rim+ lesion** | **Total** |
|---|---|---|---|
| **No disability** | 28 (70%) | 31 (55%) | 59 (61%) |
| **Disability** | 12 (30%) | 25 (45%) | 37 (39%) |
| **Total** | **40 (100%)** | **56 (100%)** | **96 (100%)** |

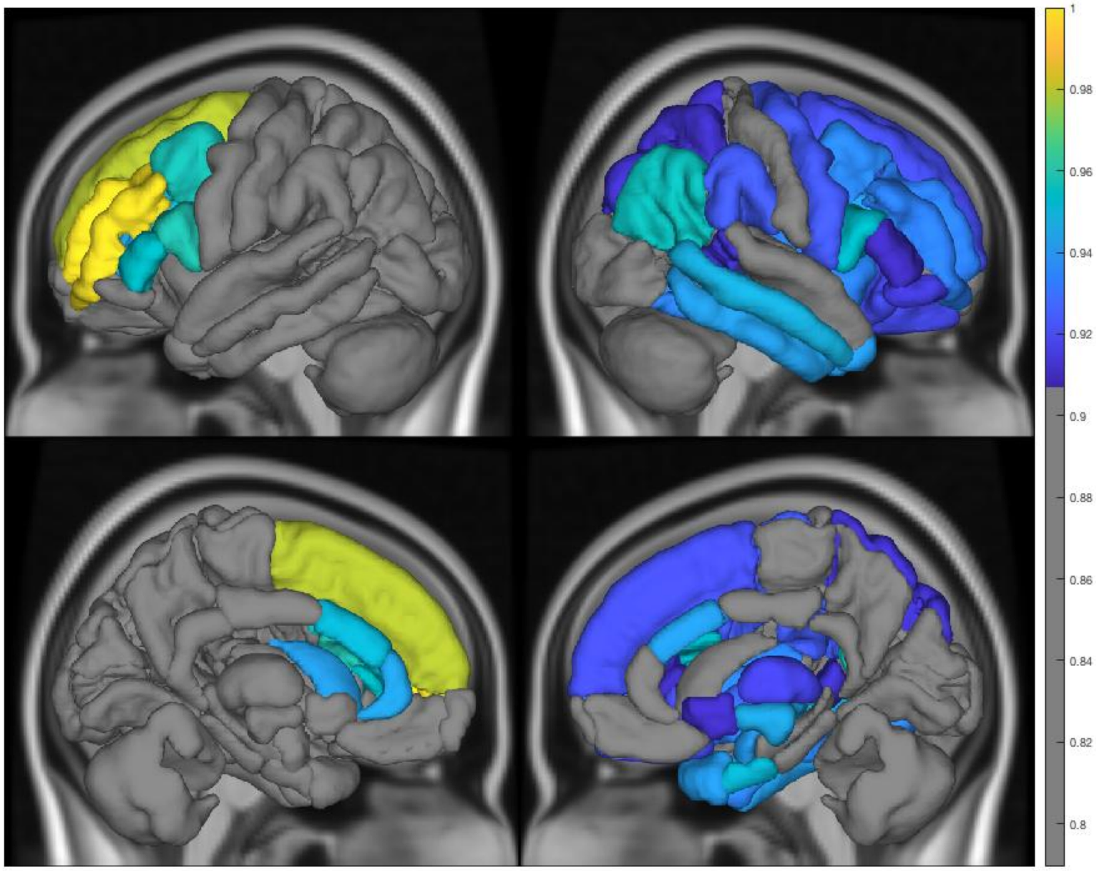


**Supplementary Figure 1:** The gray matter regions that had significantly greater (corrected $p < 0.05$) structural disconnection due to rim- lesions (larger ChaCo scores) in pwMS that had disability compared to those with no disability. The color bar represents the relative group comparison statistic obtained from Wilcoxon rank sum test.

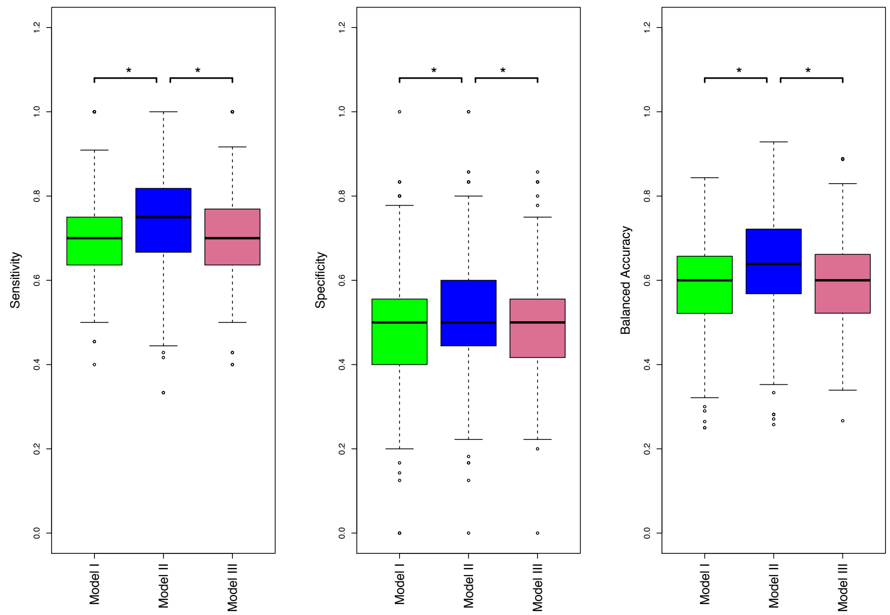


**Supplementary Figure 2:** Classification results (sensitivity, specificity, and balanced accuracy) in distinguishing pwMS according to disability group**.** *indicates a significant difference in that metric between pairs of models, corrected $p < 0.05$.
