## Supplementary material for "Structural disconnectivity from quantitative susceptibility mapping rim+ lesions is related to disability in people with multiple sclerosis": IRB approval document

Weill Cornell Medicine

Institutional Review Board

Amendment Approval

Date: 08-Dec-2020

| | |
|---|---|
| Principal Investigator: | Susan Gauthier DO |
| Submission Type: | Amendment |
| Protocol Number: | 0711009544 |
| Protocol Title: | Weill Cornell Multiple Sclerosis Database |
| Nature of Amendment: | We are keeping Neha Vijayvargiya removed from study personnel and not adding anyone new at this time. Most importantly, we would like to link an additional NIH grant to this IRB. The grant number is R01 NS102267. We are also including a more descriptive statement on future research that will be done with stored data in general study design question 4.2 . Specifically, the data will be used for a study titled "Examining the effect of Tecfidera on iron load and tissue damage in developing and established WM lesions in MS patients" with protocol number 1803019058. Subjects can be retrospectively enrolled because they have contributed all data necessary through the research repository on this study. |
| IRB Approval Date: | 08-Dec-2020 |

Dear Dr. Gauthier:

The Expedited has conducted an expedited review and approved the amendment for the abovementioned study via expedited review procedures as per 45CFR46.110.

Click here to access your submission package: Submission Page

If you have any questions, please contact the Office of Research Integrity at 646-962-8200.

Sincerely,

WCM Institutional Review Board

Please note the following important information about this approval:

- Institutional Review Board approval does not represent institutional approval. Please check the Study Activation Status Page (SASP) in wrg.weill.cornell.edu and/or ensure that all ancillary approvals are received before commencing research.
- Billing Compliance: This approval is contingent upon continued adherence with institutional billing compliance policies.
- Information about the WCM IRBs: The Weill Cornell Medicine (WCM) Institutional Review Board (IRB) is constituted as required by the Federal Office for Human Research Protections (OHRP). WCM holds a Federalwide Assurance (FWA) with OHRP. The FWA number is FWA00000093. The WCM IRBs are registered on that FWA. The registration number for the IRBs are: General IRB #1 IRB00009417, General IRB #2 IRB00009418, Cancer IRB#1 IRB00009420, Cancer IRB#2 IRB00009421, and Expedited IRB IRB00009419. Should you need additional information about the terms of the WCM FWA or the WCMC IRBs, please refer to https://research.weill.cornell.edu/institutional-review-board-irb
- ClinicalTrials.gov: **To comply with applicable requirements (HHS, NIH, and/or ICMJE), you must register prior to the first subject enrollment for studies that meet the following criteria:**
    - Interventional investigator-initiated NIH-funded WCM studies
    - Interventional investigator-initiated WCM studies evaluating at least one FDA-regulated drug, biological, or device product (except for phase 1 and small device feasibility studies)
    - Any research study that prospectively assigns human participants or groups of humans to one or more health-related interventions to evaluate the effects on health outcomes

  Updating of the record is required every 6 months, with results reporting required for some studies 12 months after study completion. with full name, CWID and phone number to set up a ClinicalTrials.gov account. More information is available at https://research.weill.cornell.edu/clinicaltrialsgov
